# Fitting the Reproduction number from UK coronavirus case data, and why it is close to 1

**DOI:** 10.1101/2021.09.23.21256065

**Authors:** Graeme J Ackland, James A Ackland, Mario Antonioletti, David J. Wallace

## Abstract

We present a method for rapid calculation of coronavirus growth rates and R-numbers tailored to the publicly available data in the UK. The R-number is derived from time-series of case data, using bespoke data processing to remove systematic and errors and stochastic fluctuations. In principle, growth rate can be obtained by differentiating the reported case numbers, but in fact daily stochastic fluctuations disqualify this method. We therefore assume that the case data comprises a smooth, underlying trend which is differentiable and a noise term. The approach produces, up-to-date estimates of the R-number throughout the period of data availability. Our method is validated against published consensus R-numbers from the UK government, and shown to produce comparable results. A significant advantage of our method is that it is stable up to the most recent data, this enables us to make R-number estimates available over two weeks ahead of the published consensus. The short-lived peaks observed in the R-number and case data cannot be explained by a well-mixed model and are suggestive of spread on a localised network. Such a localised spread model tends to give an Rt number close to 1, regardless of how large R0 is. The case-driven approach is combined with Weight-Shift-Scale (WSS) methods to monitor trends in the epidemic and for medium term predictions. Using case-fatality ratios, we create a narrative for trends in the UK epidemic increased infectiousness of the alpha and delta variants, and the effectiveness of vaccination in reducing severity of infection.

## 1 Introduction

During the coronavirus epidemic, the so-called “R-number” has become one of the best-known concepts from epidemiology. It can be defined as *“the average number of onward infections from each infected person”*. It is conventional to define *R*_0_ as the R-number at the outset of an outbreak, and *R*_*t*_ as its value some time *t* later. The significant feature is that an R-number greater than 1 implies a exponential growth in case numbers, whereas less than 1 implies exponential decay. Typically *R*_*t*_ < *R*_0_ due to acquired immunity or behavioural changes reducing spread.

### 1.1 Defining R

In a real epidemic, this conceptual definition of *R*_*t*_ is ambiguous: it may refer to people infected at time *t*; or to people infectious at time *t*; or to the rate of infection at time *t*^1^. The first two definitions incorporate infections in the future, and therefore under these definitions *R*_*t*_ is defined in that way is unknowable at time *t*. If using the third definition, the conversion from growth rate to *R* depends on some model for how the epidemic is spreading, for example “exponential growth” which is generally true only for a homogeneous, well-mixed population. If the outbreak is spreading in space, then infectors may come from a different population from the infectees, and the epidemic will be limited by diffusion and will not be exponential^2^.

Further ambiguity comes from the term “average”. This could refer to either the arithmetic or geometric mean. The average might be taken over the whole population, or only over those who are actually infected.

Any epidemic model which does not represent each individual person cannot simply count the number of subsequent infections per person. Thus definitions of *R* are usually related to growth rate. Assuming that the of number of new infections is proportional to the number of currently infected people, the growth rate is:

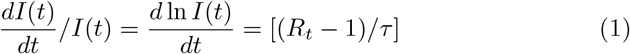

which introduces a timescale *τ*, similar to the time between infections and therefore referred to as the generation time^3^. This definition of *R*_*t*_ shares important features with other definitions, in particular that *R* = 1 is the critical value separating a growing and diminishing outbreak. Its advantage is that it does not depend on future events.

Our approach to *R* is even more pragmatic. We define *R* as a quantity based directly on available data which satisfies the constraint that *R* = 1 is the critical value and reproduces the rate of growth of the epidemic. In practice, this means using the equivalent to equation 1 using reported positive PCR test data (cases, C(t)) in place of infection data. This leads to a different *R*_*t*_, defined by

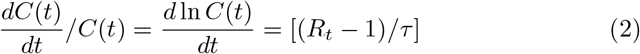

Not all infections will be reported, and reported cases may include false positives. A vital feature of this equation is that even if only a fraction of infections are reported, that fraction cancels out: *R*_*t*_ is independent of systematic underreporting.

### 1.2 Using *R* derived from case data for policy

The *R* number is often used by policymakers to trigger interventions. It is particularly useful because it is a **leading indicator**: *R* can been seen to exceed the epidemic value *R* = 1 long before cases, hospitalizations and deaths reach critical levels. Local measures of *R* enable governments to use well-focused interventions to achieve maximum disease suppression with minimal disruption. However, care must be taken that the correct measure is being used.

The growth rate is determined by infectees, but many policies are aimed at infectors. If these groups are different, ignoring this distinction can lead to misapprehensions. For example, a rural area may have *R* < 1 such that cases are entirely driven by incomers from an urban area. If the I(t) in equation 1 is dominated by incomers, the *R* value calculated from equation 2 will reflect the *R* of the urban area, and unaffected by local measures for the suppression of *R*^4^

To illustrate the effect of mixing on R, we examine a two population SIR model. Consider an urban population 1 which lives mainly in a high R-area (*R*_1_ = 2), the rural population 2 which lives mainly in a low R-area (*R*_2_ = 0.5). *R*_1_ and *R*_2_ follow the normal definition of *R* within the SIR model based on contact between individuals. For simplicity, we assume the populations are of equal size. The urban population spends some fraction *x* of its time in the rural area.

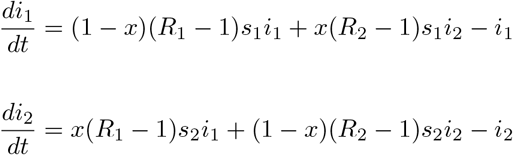

where the populations *s, i, r* are fractions of the total, and 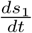 and 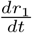 follow trivially from the terms in 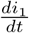.

Assuming that the measurable quantity is the number of cases *i*(*t*), figure 1 shows the results of applying equation 1 to infer *R*_*t*_. Two cases are considered:

**Figure 1:**
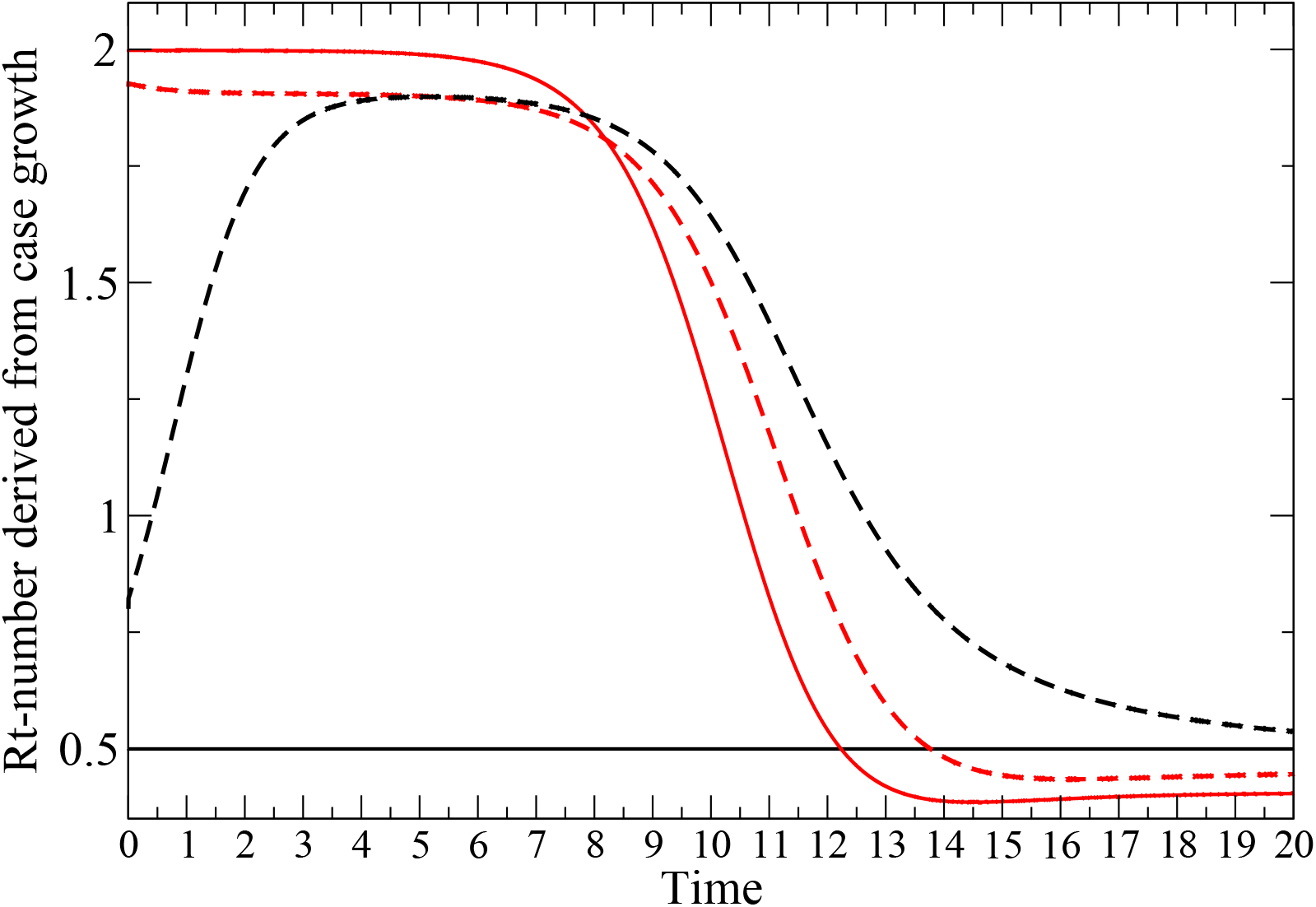
Detectable R-numbers from coupled SIR model. Both i(0)=0.00001. Red lines are Rt derived from population *i*_1_, black lines relate to populations *i*_2_ Solid lines show no mixing (*x* = 0), dashed lines have (*x* = 0.05). The time axis is in units of the generation time.

With x=0, the populations are unmixed: in the urban population *s*_1_ is reduced until herd immunity is obtained; in the rural population the *s*_2_ ≈ 1 and initial infection dies out exponentially.

A modest 5% mixing is enough to change the picture completely: rural case numbers are driven by incomers, and the measured R-numbers of the two regions become equal.

Although framed in terms of geographical populations, the same principles apply to any coupled subsystems with different levels of infection, e.g. age groups, vaccinated/unvaccinated, isolating/non-isolating. The R-number inferred from cases will always be due to the subpopulation with more cases, not the subpopulation being measured.

The relevance from a policy viewpoint is that measures imposed to suppress *R*_2_ serve no purpose in suppressing the epidemic, despite the inferred *R* in that region being well above 1.

Infection rates are also linked to seasonality, and even to weather - contact rates will change if people spend more time indoors. This is a real effect, but weather-induced fluctuation will be high-frequency and indistinguishable from noise. Policy should not depend on previous weather, so ignoring weather as a factor is appropriate.

### 1.3 R in the UK

Here, we attempt to define and model the *R*-number in the UK based on contemporary data. We constrain ourselves to use only publicly available data, in particular the case data from the government website[1] and similar data from the Scottish government[2]. Our approach is pragmatic - we do not assume it to be in any sense “more accurate” than other authors: indeed, our expectation is that it will be less accurate due to the data constraints and the simplicity and transparency of the model. *“An 80% right paper before a policy decision is made it is worth ten 95% right papers afterwards, provided the methodological limitations imposed by doing it fast are made clear*.*”[3]*

Our model provides *R* estimates some two weeks ahead of those published by the UK government[1]. To examine whether our method is at least “80% right” we will benchmark our predictions by hindcasting against “gold standard” model-based work contributing to government policy.

In most epidemic theory models, *R*_*t*_ is uniquely defined by the rate of growth of the number of infections of the epidemic.

In the second wave we had reasonable data for the number of cases as a function of time from positive PCR tests *C*(*t*). This is significantly less than the number of infections, as measured by the Zoe and ONS random/weighted cohort survey *I*(*t*), but they they are proportional which, as already discussed, is sufficient for *R* calculation. There is also a delay between infection and test of approximately 5 days, such that any estimate based on case data will be out of date. We use reports of the first positive test based on PCR by sample date - the ONS cohort survey typically has a larger time between infection and reporting, so is less useful for up-to-date surveillance.

Using case data rather than the cohort study introduces an important bias, towards a group which has above average level of infection. If one imagines every individual has their own *R*-number, then the measured *R*-number is not the average of those. This is because the people with higher individual *R*-numbers are more likely to be infected, and therefore more likely to be included. As an example, consider two unconnected cities with *R*-numbers 1.5 and 0.5 respectively - only the first suffers an epidemic and contributes to measured cases. Thus the measured average *R*-number across the two cities is 1.5.

Another important issue is that because growth is exponential, removing noise using simple averages of *R* can be misleading. As an example, suppose the “true” *R* rate across two generations is 1, such that the third generation has as many cases as the first. Now, suppose due to noise the measured *R* rates are 2 and 0.5 such that, again, the third generation has the same number of cases. This is all consistent, but applying the average *R* (1.25) would wrongly suggest a 56% increase. A harmonic mean gives a correct result. In general, using geometrically averaged *R* numbers in place of noisy data always implies more cases than are present in the data.

## 2 Methods

### 2.1 Inference from the future and the Second Law

We have previously used a case-driven kernel compartment model (WSS) to track the course of the epidemic. In this method, each reported cases on day t generates a probability distribution into other compartments in the future. The simplest case is a two-compartment model with compartments being cases C(t) and deaths D(t), in which we write:

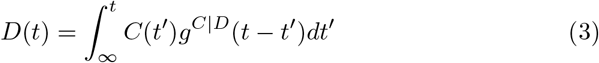

In which *g*^*C*|*D*^(*t*) is the distribution of times between reported case (i.e. positive test) and death, has been measured from case data to follow a Gamma distribution[4] this can be written as the probability of death on day *t* given case on day *t′ p*(*D*(*t*)|*C*(*t′*)).

We will be using case data to infer *R*. There is some distribution of times between infection, symptoms appearing and positive testing. It may appear that one could apply Bayes theorem using the “probability that infection occurred on day *t*, given a positive test on day *t* + *t′*” to infer infections from the case data.

However, to do so violates an even more fundamental principle - the Second Law of Thermodynamics - the relevant form of which states that for an irreversible process, entropy (in this case uncertainty about dates) must increase.

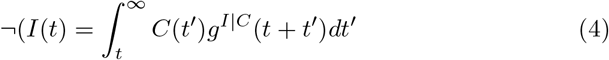

In the present case, a sharp rise in *R* (e.g. from lifting of restrictions) will produce an increase in cases spread across several days. Following the second law, the sharp feature should precede the broad one.

But by projecting the *case data* backwards in time by applying the distribution of time lags, the features in *R* will be spread out, giving an implausible situation where sharp features in the case data arise from slow changes in *R*.

Thus *R* defined on cases (eq. 2) will be more slowly-varying than *R* defined from infections (eq. 1).

We note that equation 4 can be read as “Bayes theorem cannot be applied backwards in time to an irreversible process”. The problem lies in assuming that *g*^*C*|*I*^ (*t* − *t′*) is independent of *t′*. In some previous work the “reducing entropy” problem is avoided by using strong low-entropy priors for the infection-based R, e.g. insisting that it is piecewise constant[5].

### 2.2 What is the case data?

The UK case data (Fig. 2) entails daily reports of order 10^4^ positive tests. We assume this will be subject to day-to-day statistical stochastic noise^5^ 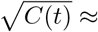 100, and variations in reporting depending on day of the week so we write the raw data as:

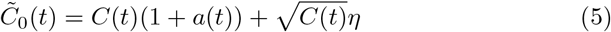

**Figure 2:**
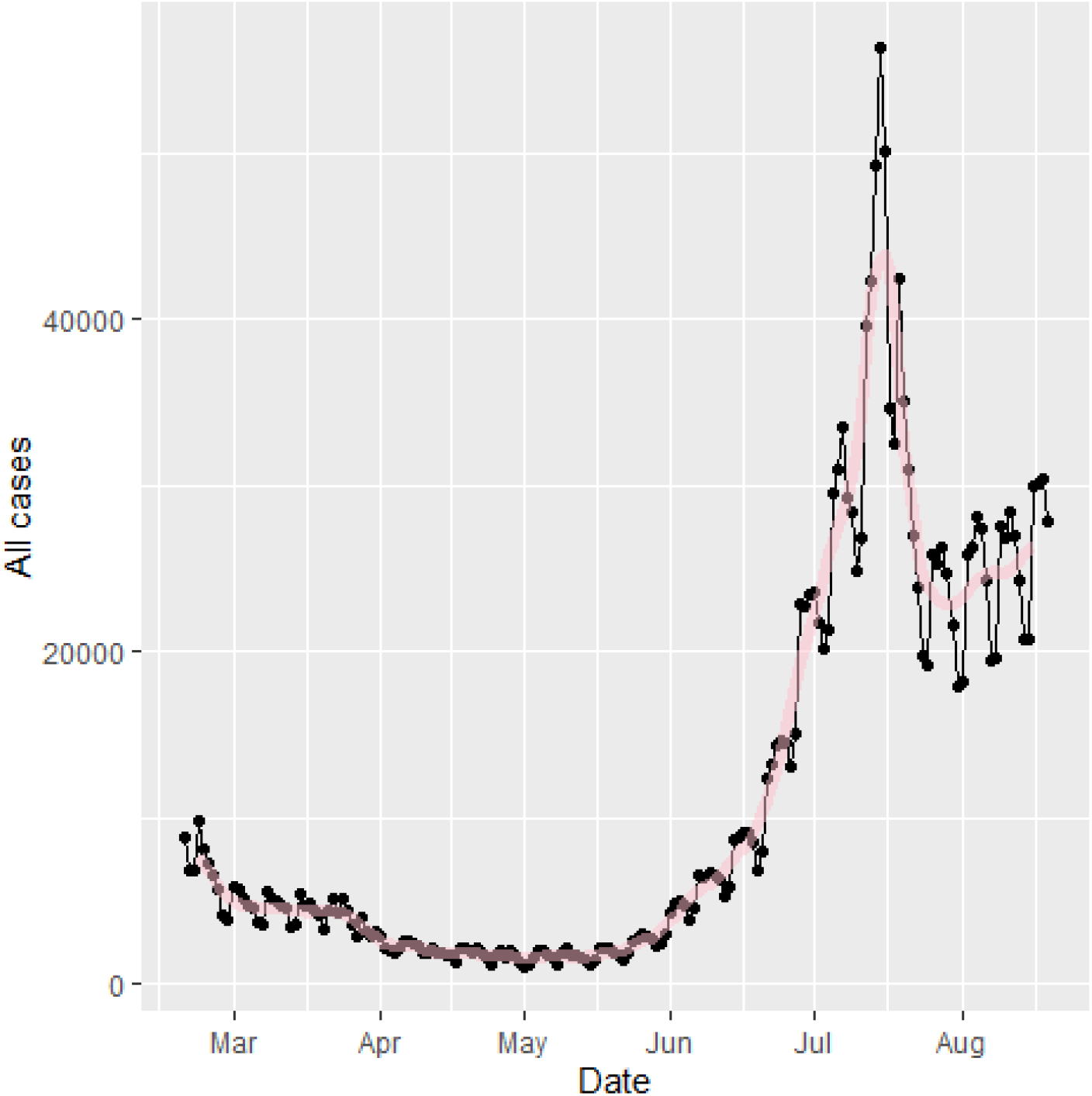
Raw official data for cases for England. A strong weekly oscillation is evident. Although it is plausible that more infections happen on weekdays when people are at work, we will assume the oscillation is from the amount of testing

Where 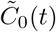 is the reported data, defined only at integer *t, C*(*t*) is the underlying trend, *a*(*t*) is a systematic reporting error and *η* is a random variable. *C*(*t*) is a differentiable function, but *η* is not. To differentiate this function requires methods from stochastic calculus, but for practical purposes we move directly to algorithms to deal with the data. In practice, we shall require that the *R*-number be defined in such a way that, if we re-create the epidemic by integrating *R* through time, it must reproduce the actual epidemic size.

### 2.3 Identifying and eliminating the systematic errors, *a*(*t*)

We identify four courses of systematic error in the data:

- False Positives and Negatives
- Underreporting at weekends, and associated catch-up
- Underreporting on holiday, and associated catch-up
- Delayed reporting at the end of the time series
- Misreporting ^6^

The false positive rate was identified from the case fatality rate (CFR)[6] as a significant fraction of reported cases in September 2021, without which the CFR would have been unreasonably low. It was estimated to be approximately 0.4%. This is much higher than previously assumed[7] based on the *total* fatality rate in the summer. The cause of this discrepancy may be cross-contamination[8]. The amount of testing, and by implication the number of false positives, has varied relatively slowly compared with the changes in *C*(*t*).

In terms of *R* number calculations, the false positive rate, as with anything which means that a constant fraction of cases are not reported, has no effect.

Systematic underreporting of cases at the weekend is evident in the data.It is systematic, so we cannot treat as an enhanced stochastic term. To eliminate this, we make an assumption that across the epidemic the number of infections is independent of the day of the week. Specifically, we rescale the cases by a factor:

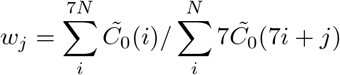

With *N* the number of weeks of data. This means that the total number of cases on Mondays is reset to be equal to the total on Sundays, etc. It removes an obvious source of systematic error.

Across the Christmas period the weekend effect breaks down, and there are even larger fluctuations in the case data. Hence, over the 12 day period from day 153-164 (Dec 24th-Jan 4th) we fit a straight line through the cases data, constrained to preserve the total number of cases.

We also investigated a rolling 7-day average. This gives some smoothing, but systematically flattens peaks and fills troughs in the data. The calculation was also repeated by taking seven separate streams of data, one for each day, calculating *R* based on seven day changes, then averaging these values.

There is a short delay between positive test and reporting. Using historical data we found this to be systematic, which allows us to make even more up-to-date measurements. Within Scotland, we find ratios between cases reported for the three most recent days and the final totals for those days. These are of 2.9 (±0.2), 1.05 (± 0.01), and 1.005(± 0.002) respectively.

The data with these time-dependent systematic errors removed is shown in Fig 9 denoted by:

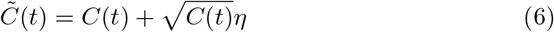

Henceforth, we will use this 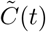 as case data.

### 2.4 Stochastic Differentials

If we had a differentiable *C*(*t*), we could evaluate *R* as defined in equation 2. Unfortunately, the data is 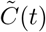, not *C*(*t*) - it is only defined at integer t and the stochastic noise is still present. Nevertheless, we can integrate the equivalent of Eq, 2 and calculate 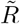, the “R-number with stochastic noise”.

We make a further assumption that *R*_*t*_ and *τ* are slowly varying in time, so that we can ignore their time-derivatives. Integrating equation 2, we find that

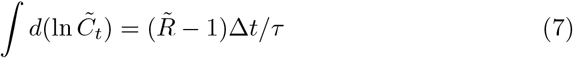

To perform the integral we should use stochastic calculus, and this introduces some ambiguity: Case data is available daily, so we can take the discretized form of this equation using the Stratonovitch form:

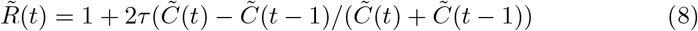

Or its Ito Calculus equivalent

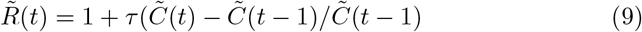

Alternately, we can define 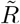 from the exponential form

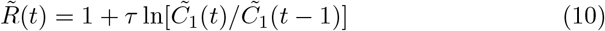

The data is discretised with a step of one day: we also investigated eliminating weekend effects by increasing the step to seven days

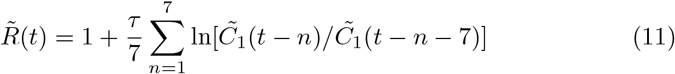

In each case, we write 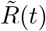, noting that 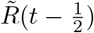 is more appropriate. All the approaches above were tried, and terms of final results for *R*, we found little difference between any of these methods. However, if one attempts to regenerate the *C*(*t*) using these 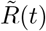 by integrating eq.2, then only the exponential discretisation (10) reproduces the time series correctly.

### 2.5 Estimating the uncertainty in R

However *R* is calculated, it involves sampling noisy data over some time, during which *C*(*t*) itself is varying. Early models assumed that *R* is constant between changes in policy interventions[5]. If true, this assumption would allow the fitting errors to be calculated precisely, but there is strong evidence that *R*(*t*) varies steadily in time due to varying compliance, increased post-infection and post-vaccination immunity and the rise of variants. If *R* is varying in time, there is a conflict between reducing the stochastic error by sampling over many days, and having an up-to-date estimate. We postulate that *R*(*t*) is not only differentiable, but also all its derivatives are slowly varying in time. This means that we can reduce uncertainty and make more up-to-date measurements of *R* by estimating 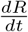 and higher derivatives, which is best done using some smoothing function (see section 2.8).

Since *C*(*t*) grows exponentially with *R*(*t*), it will be more rapidly varying, and because of variable time from Infection to Testing, *I*(*t*) will vary even more rapidly. The case data actually defines a growth rate, which is dedimensionalised by the generation time *τ*. Our calculated (*R* − 1) is directly proportional to *τ* and so probably the largest uncertainty in *R* come from the uncertainty of *τ*. We take a value of *τ* = 5 days.

### 2.6 Do cases rise exponentially?

While R is a well-defined concept in terms of onward infections, the idea of “Growth rate” assumes an exponential process. We consider three models for predicting the case data C(t):

- Same as yesterday: *C*(*t*) = *C*(*t* − 1)
- Exponential Growth *C*(*t*) = *C*(*t* − 1) ln[*C*(*t* − 1)*/C*(*t* − 2)]
- Linear Growth *C*(*t*) = 2*C*(*t* − 1) − *C*(*t* − 2)

Averaged across all UK regions, we find that “Same as Yesterday” gives the smallest RMS and mean absolute error, with linear growth about 1% better than logarithmic. The effects of noise are significant, but there is no evidence that logarithmic growth gives the best short-term prediction of growth.

### 2.7 What value for R causes an epidemic?

In the SIR model, we have exponential growth and any value for *R*_0_ greater than 1 causes an epidemic in which a finite fraction of the population becomes infected. The ODE approach to SIR assumes complete mixing of the population, but network effects[9, 10, 11, 12] can significantly raise the threshold for R to cause an epidemic. The exact form of the UK contact network is not known, but there are some well-defined mathematical approximations which can be implemented in an autonome-based model, and it has long been known that allowing spatial variation can affect behaviour in many contexts[13, 14, 15].

We simulated a stochastic agent-based model of SIR-autonomes^7^ with different types of connectivity:

- Random connections on a fixed network
- Regular lattices (square, triangular, cubic)
- Small world lattices, with random long-ranged connections added to a regular lattice

It is natural to interpret the lattice as a division of people in space, with contact most likely with those living nearby. However, other interpretations are possible, for example age group: the POLYMOD[16] shows that contact is primarily with people in one’s own age group.

Each simulation is seeded with 10 infected sites, and transitions from *S* → *I* or *I* → *R* are implemented according to the Gillespie algorithm[17]. Once the network is defined, this model has only one parameter, *R*_0_, the ratio of attempted infection rates ^8^ to recovery rate.

It is evident from Fig.3 that *R*_0_ = 1 is a poor predictor of whether the infection triggers an epidemic. The ODE result of a threshhold at *R*_0_ = 1 is recovered for a fully connected network. Less densely connected random networks require dozens of connections per node to generate an epidemic with *R*_0_ = 1. For the sparser networks the total number of infections can be significantly less than the total population. For two dimensional lattice networks the threshhold for an epidemic is *R*_0_ = 2. This can be understood by noting that the SIR-lattice can be mapped to a reaction-diffusion equation, which generates a travelling wave[18] moving at constant velocity - in the SIR context autonome behind the wave are predominantly I and R, while ahead of the wave they are S. New infectees typically lie on the boundary between previously infected and fully susceptible regions - so from early in the epidemic approximately half their neighbours are S.

**Figure 3:**
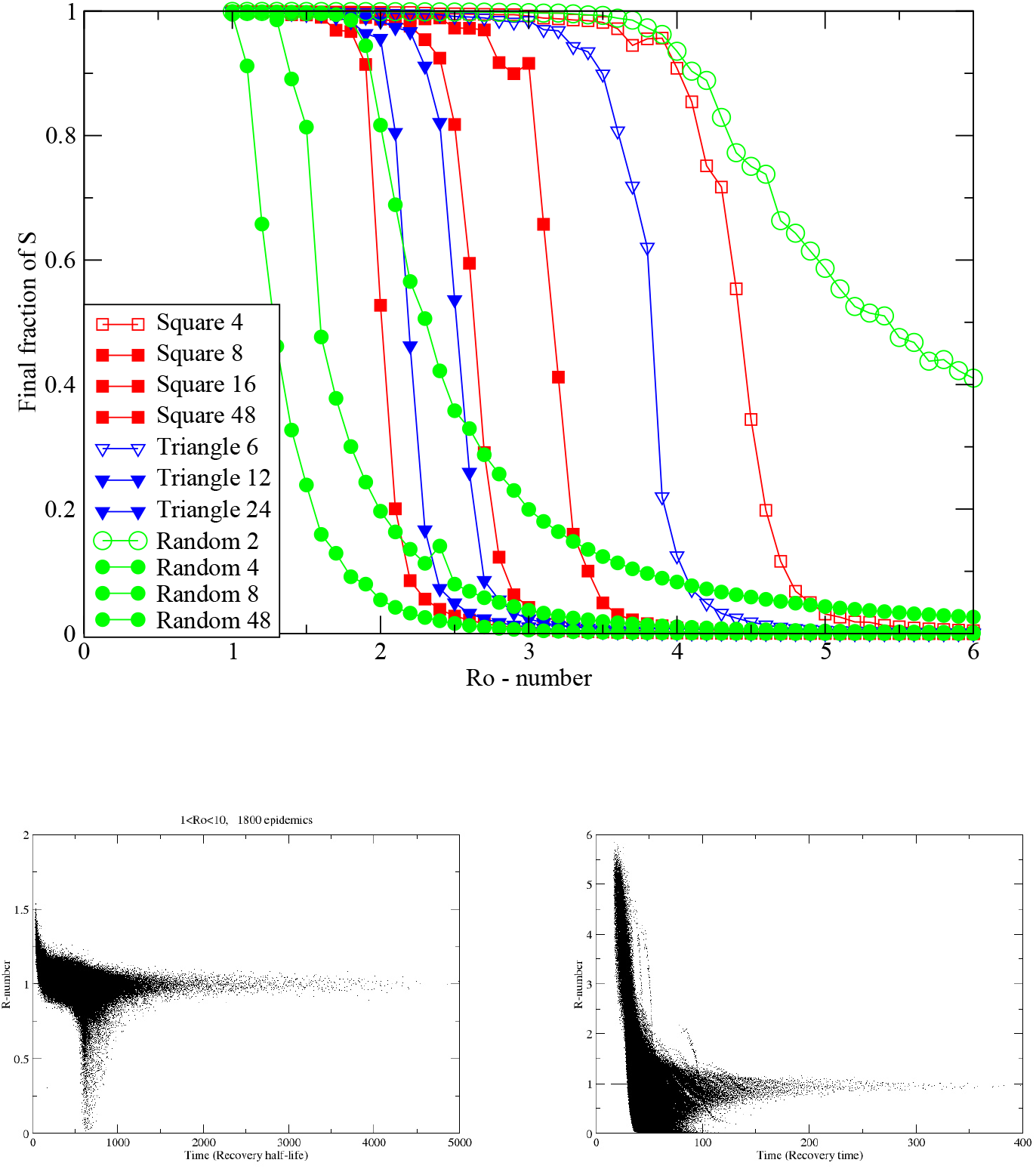
Figure shows (a) the size of the final epidemic for various network structures and values of *R*_0_. Legend gives the different lattice structures and the number of connections each has. To obtain the given *R*_0_, infection probability per link is lower in more highly connected lattices. Epidemics spread on the two dimensional lattice only for *R*_0_ >, the smallest values appearing for the densest networks. (b) Scatterplot of measured *R*(*t*) = −∆*S/*∆*R* from simulations with 8-neighbour square lattice, 500,000 sites, *R*_0_ ranging from 1 to 10. 20 simulations at each 0.1 increment in *R*_0_ are shown. Timescale has recovery rate set to 1. Other lattices are similar. (c) Small-World version of (b) with 8 neighbours plus one added long range connection.

The R-number for these lattice models is shown in Fig.3b,c). These scatterplots come from many hundreds of different simulated epidemics with an order of magnitude variation in 1 < *R*_0_ < 10. Each point represents the value of *R*_*t*_ which would be measured in the epidemic. Individual epidemics are not tracked, but two distinct behaviours are evident: either the epidemic does spread, and *R*_*t*_ drops to zero after some time, or it does spread across the system.

Remarkably, for any *R*_0_ large enough to generate an epidemic, *R*_*t*_ tends to 1, after some transient time. This is completely different from an ODE-based well-mixed SIR model, for which the value of *R*_*t*_ decreases steadily with time with no special behaviour as it passes through 1 (Fig.1).

The lattice model neglects long range connections: we introduce these with a “Small World” network in which additional random connections to anywhere in the system are added to the 8 neighbours. The R-numbers for such a network with one long-range connection per site are depicted in Fig.3c). The plot is broadly similar to Fig.3b), although notice the difference in the axes. We see that

- The timescale of the epidemic is very much reduced by the long range connections.
- The high *R*_0_ epidemics retain a high value of R because the epidemic is over before the transient ends
- Intermediate values of *R*_0_ still tend to *R*(*t*) = 1

### 2.8 Smoothing the data

This 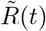 is the required *R*(*t*), plus a term arising from the stochastic noise.

Within the UK, case numbers are typically of order 10000, so we can expect stochastic noise of 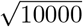, i.e. about ± 1% error in daily growth rate (which is typically of order 1%). Thus we can expect that direct calculation of growth from single day’s change, even with systematic error removed, will have 100% uncertainty. Fig 4 shows that the noise is indeed dominant, and across the pandemic the standard deviation of 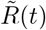 is about 0.6. This value is confounded by the actual RMS variation 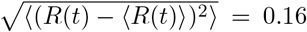, and by any slow-varying systematic errors such as the effectiveness amount of testing.. We now make our final approximation, smoothing the data to eliminate the high-frequency noise in 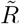 while retaining the smoothly varying signal *R*(*t*).

**Figure 4:**
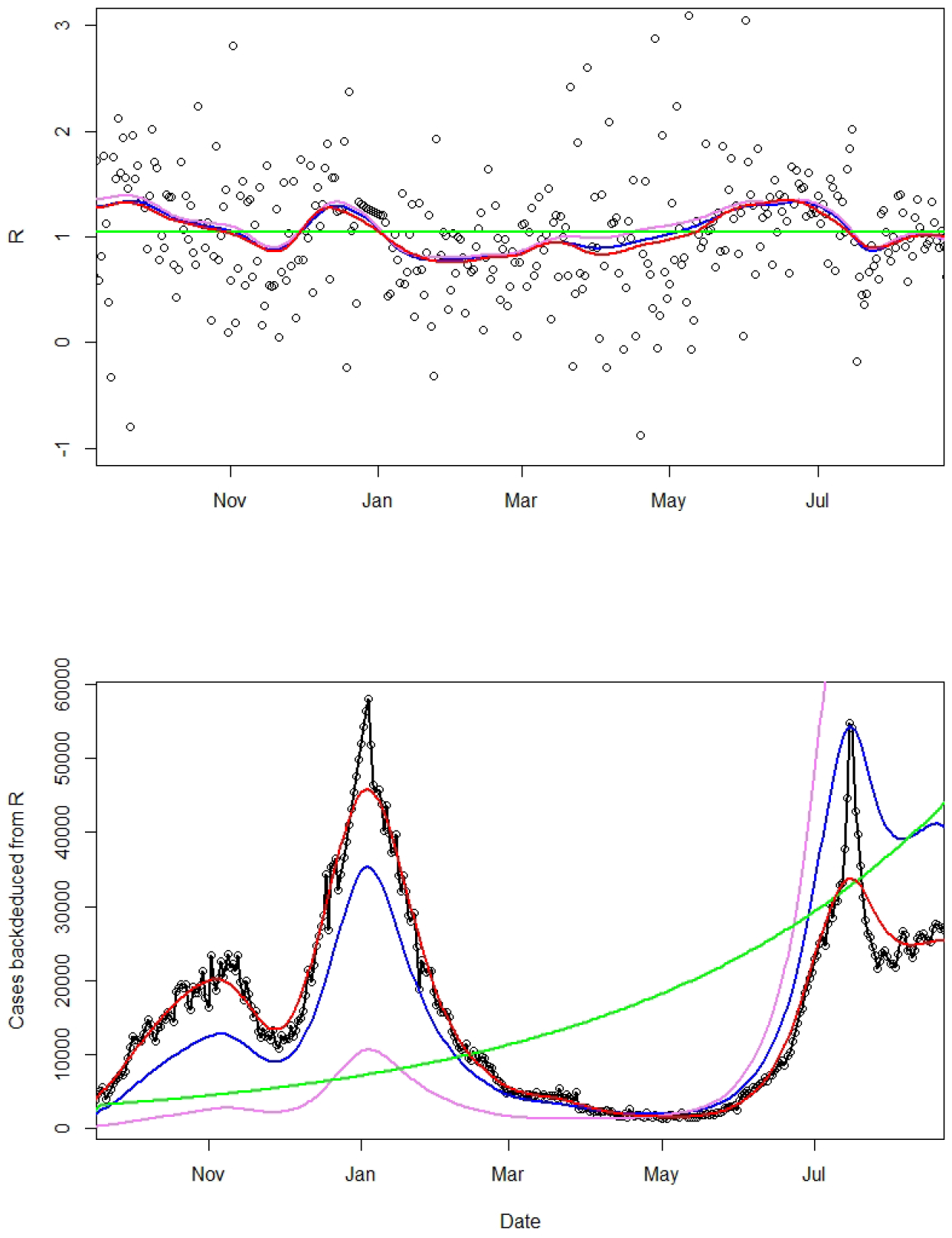
(top) *R* values calculated from various methods. Points: 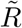 eq.10, Violet: Ito integration +smoothing of 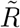, Blue: log integration +smoothing of 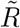, Red: smoothing of 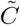 + log integration, Green: average value. (bottom) Modelled case numbers using these R-numbers from October 2020, with initial case numbers chosen to give correct total number of cases. Black circles show actual data, which 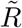. reproduces by construction.

Various standard methods of smoothing the data were considered: weekly averaging, LOESS, spline fits with various number of splines, and independent spline fits starting and finishing at, or 5 days after, imposition or removal of lockdown, to account for discontinuity in *R*(*t*) when policy changes. Where case numbers are low, the stochastic term is larger relative to the signal, so all fits are weighted the square root of the number of cases.

All smoothing methods give similar results, so we chose to use splines, and applied them to the various methods of evaluating *R*: eq.9; eq. 10; mean *R* across the entire period (1.04); C(t) from eq.6 by smoothing 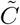. Figure 4 shows that all smoothing methods appear to give similar variation in R. However, if one attempts to reproduce the trajectory of the case numbers using these different measures small differences in *R* are magnified.

## 3 Validation

### 3.1 Sensitivity of *R* of R to fitting methods

In addition to the type of smoothing applied, the amount of smoothing leads to variations in predicted R. Fig.5 shows independent piecewise fits to periods between lockdowns and unlockings. Curiously, the discontinuous piecewise fits are found to still give a nearly continuous behaviour, the one exception being around Christmas and New Year where the reporting data is erratic and does not follow the weekly variations. So, we can reasonably assume that *R*(*t*) is a slowly varying function, and that *dR/dt* is a continuous function which can be used to improve the estimate of *R*(*t*) beyond the average over the smoothing period and into the future. All of these features mean that the uncertainty in our *R*(*t*) will be much lower that the residuals typically calculated by fitting codes, although without knowing exact functional forms, it is impossible know by how much.

A final check on the uncertainties in the method comes from comparing the R-value obtained by the different smoothing methods, and different smoothing periods (Figure 5). Reassuringly, these are all consistent within ±0.1.

**Figure 5:**
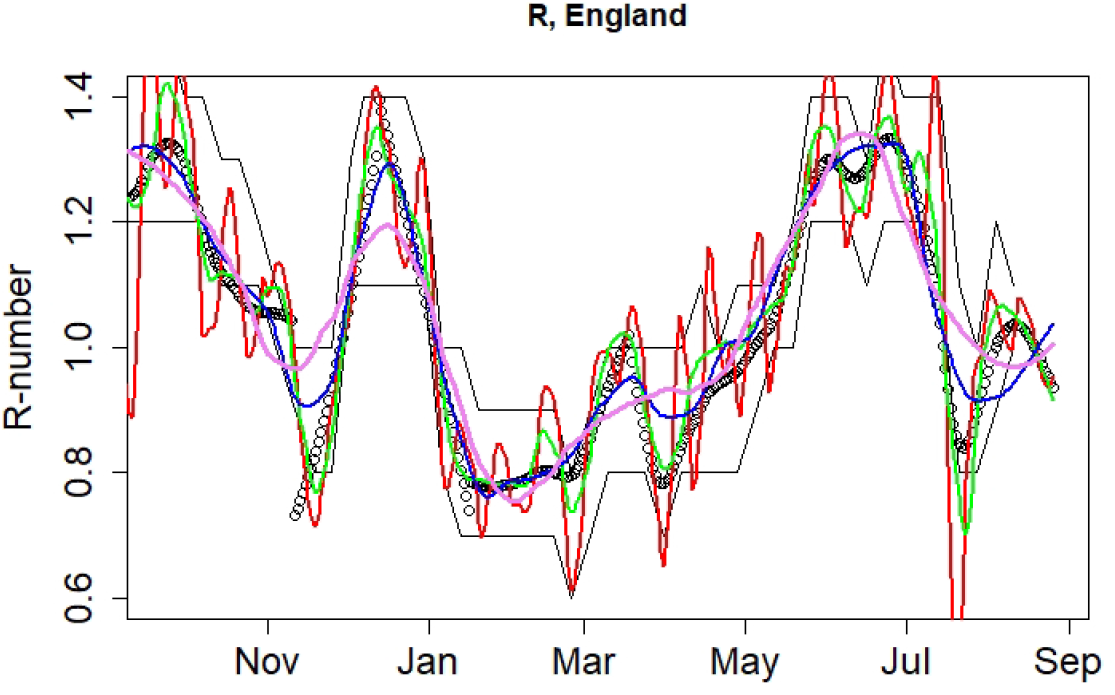
R-numbers for England. Black points are our central estimate, based on piecewise fit between major locking and unlocking events. Coloured lines (red, green, blue, violet) show LOESS smoothed R-numbers from Eq.10 with span= 0.05, 0.1, 0.2 and 0.3 respectively Black lines are the published bounds on *R* data from SPI-M consensus: to obtain this agreement, the consensus values are assigned to a date 16 days before publication.

### 3.2 Validation by Reverse-engineering the epidemic

Since the R-number is the gradient of the case numbers, it should be possible to recreate the case number data using only the R-number and the initial caseload. If 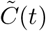 were a continuous variable, this would be straightforward, but if we smooth 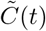 or 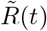 then we lose information because the smoothing process is not reversible.

Figure 4 compares with the actual number of cases with those regenerated from R-numbers. One sensible constraint is that, whatever we do, the integral on C(t) - i.e. the total number of cases - should be correct.

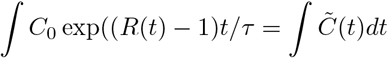

This is done by adjusting *C*_0_, the initial number of cases, which allows recreated trajectories from different smoothing methods to be compared on an equal footing.

Using 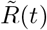 exactly reproduces the data, but all smoothed versions of 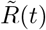 overestimate the growth rate - as evidenced by the second peak being much higher than the first when the total number of cases is set equal. Smoothing the case data first, then calculating *R* from smoothed case data, gives a better fit, with the feature somewhat broadened for reasons similar to Section4.

We note that the error is in part because *the form of the noise is not known*. So for example, if we assume a form for *η* such as white noise or Gaussian random variable such that:

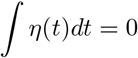

Then it trivially follows that

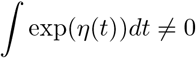

Since the *R*-number appears in the exponential of the epidemic growth, it follows that the “noise” gives a non-zero contribution to the growth rate which should or should not be incorporated in *R*(*t*), depending on *R*’s precise definition.

We see that stochastic integration using Ito’s method gives the worst results, giving a systematic overestimate of *R* which equates to too-high case number at long times. Integration using log cases performs better. The better reproduction of the epidemic suggests that it is better to treat the noise in 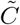 rather than 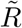

### 3.3 Validation by appeal to authority

*R* is not directly measurable, so there is no way to empirically validate these results. We therefore compare our predictions to those from more sophisticated epidemic models from the UK government’s SPI-M[19] committee ^9^. UK Government data about *R* is derived as a weekly consensus across many different methodologies and groups. ^10^

It is clear from the figure that our *R*-estimates are compatible with the reference values published 16 days later. This is reasonable, since published data is stated as being an average over the preceding weeks. The SPI-M consensus is reached during the week prior to publication in advance of the published data and are therefore available earlier to policymakers. Nevertheless, our direct method is capable of providing reliable values well in advance of the currently-published values.

A definitive empirical measurement of R is lacking, so it is possible that both simple and detailed models are similarly wrong. Regardless, our method has been demonstrated to be an excellent predictor of future published results.

## 4 Implementation and Results

### 4.1 Code and subdivisions

*R*-calculation is implemented by the WSS[20] codebase, which is publicly available and written in R. WSS use imported data updated daily, and executes within minutes on a single processor.

The WSS code generates *R*-numbers at the regional level (Fig. 6). The statistics for UK nations and nine English regions are sufficient, and are consistent with the SPI-M published values (subject to 16 day lags). We also evaluated *R* at the level of individual health boards in Scotland. These showed plausible trends, except for the smallest boards. The issue there due to insufficent data, but the fact that rural “R” values may be driven by incomers as already discussed in Sec. 11.2 means that case-data may not be indicative of community infection rates in those areas. The local authority regions in England also often have too small data for accurate evaluation, although a combination of large R and high case numbers can be indicative of local hotspots or superspreading events.

**Figure 6:**
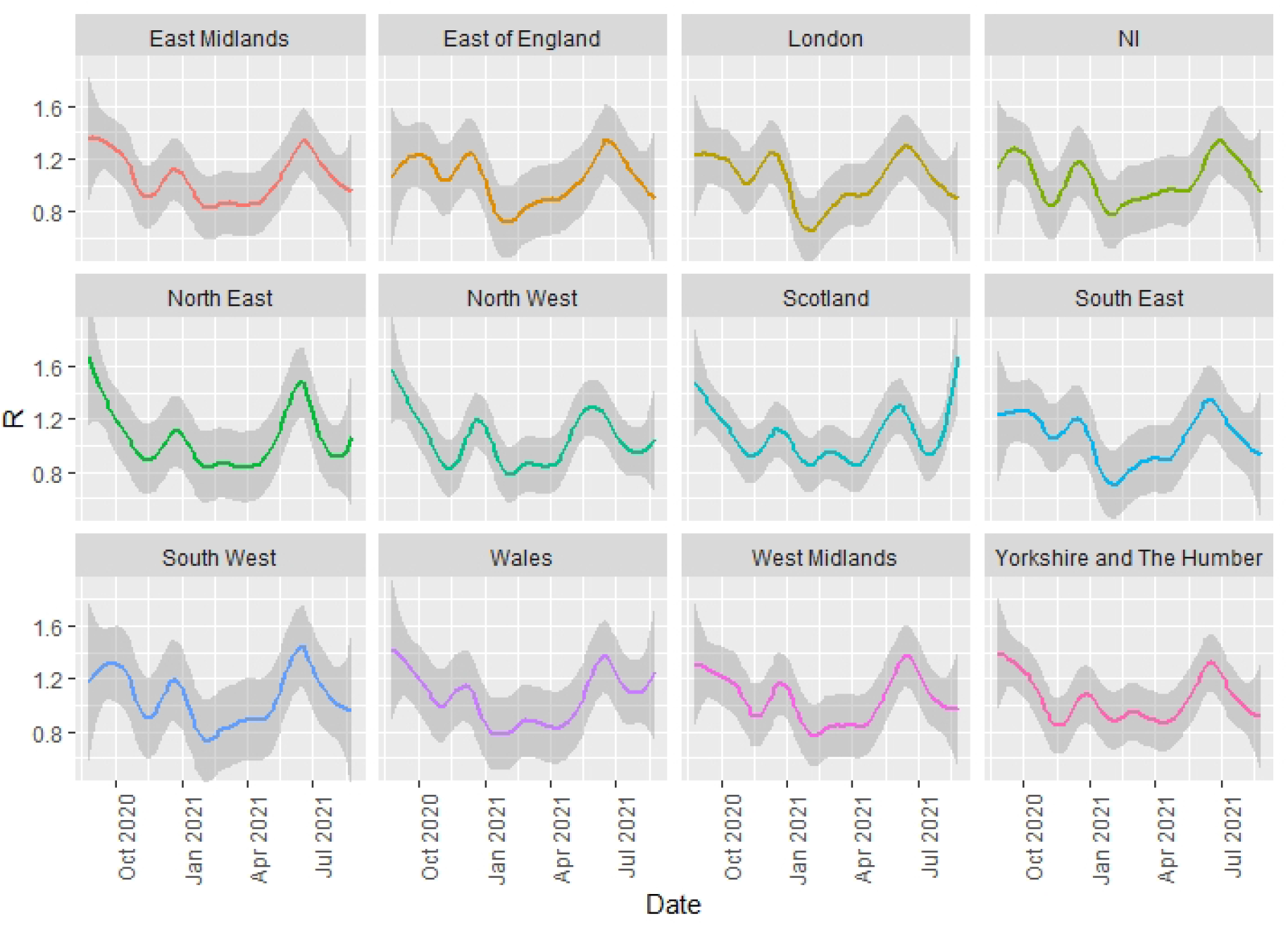
R-numbers for UK nations and English regions, shading is the mean squared error associated with the smoothing (here, LOESS with span = 0.3). December and June peaks associated with the alpha and delta variants are evident in all regions. Blips in September and March correspond to low case numbers, and may be artifacts.

**Figure 7:**
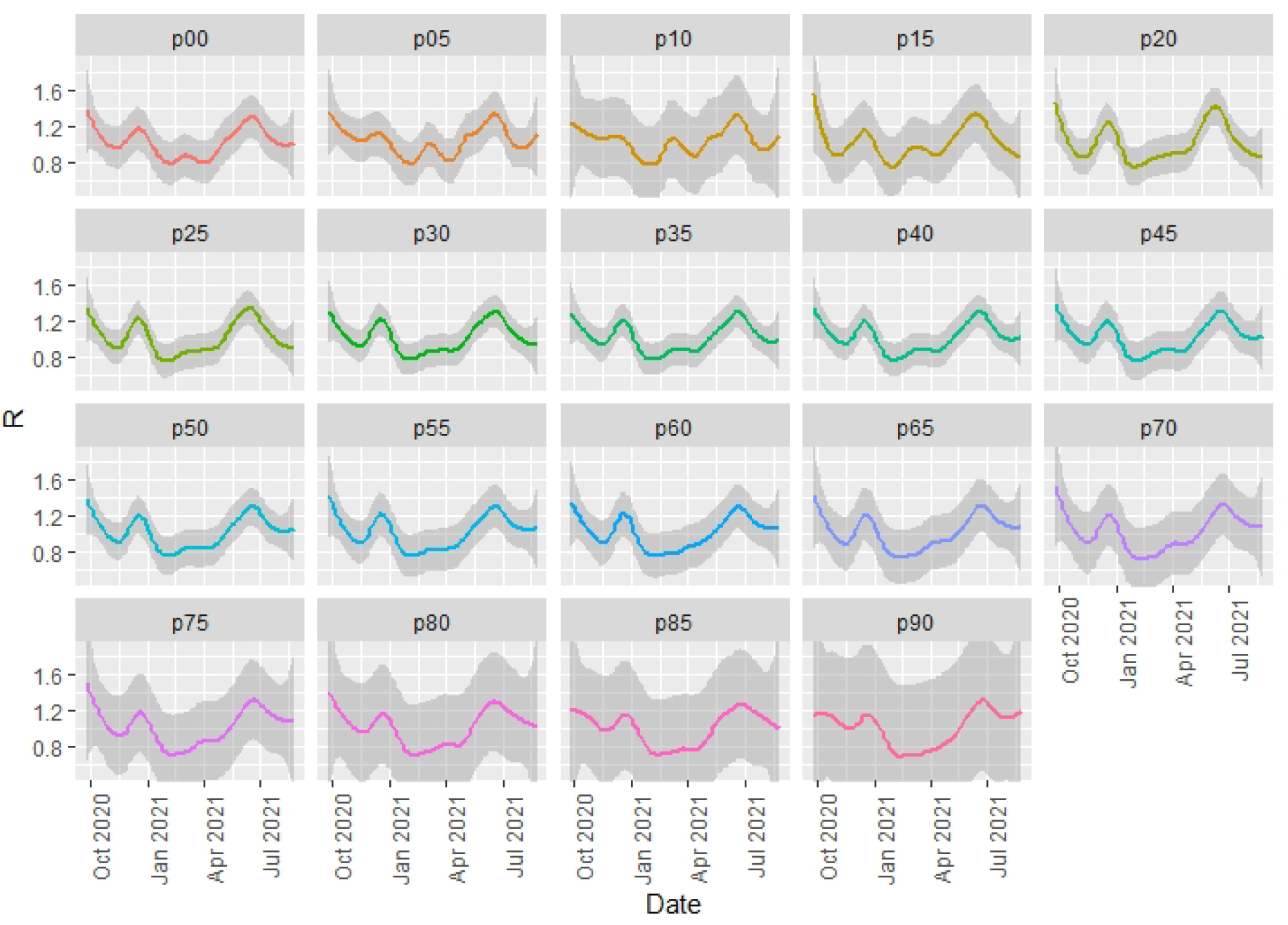
WSS R-number prediction method applied to case data split by five-year age group, p indicating the youngest yeargroup. Data is averaged across all England, smoothed using LOESS span=0.3, shading is uncertainty based on local fit. See main text explaining why this is a scaled growth rate and not a conventional R-number.

The data can also be sliced to provides a growth-rate breakdown by age group (Fig.6). Breakdown by age has a similar problem to regions because case data refers to infectees, not infectors - and generally infectors are in a different age group from infectees[16]. This intergenerational mixing is particularly true for families, hospital and carehome situation. Specifically, when case numbers are unevenly distributed across age groups, the “R-numbers” ascribed to older age groups do not imply that these people are responsible for infection.

### 4.2 Vaccination effects

Vaccination is known to reduce transmissibility of the virus by 60-90%. It may seem mysterious that there is little sign of an effect of vaccination in the national or regional R. To understand this, one needs to look more deeply into the data. Fig. **??**, the “R-number” sliced by age group, shows the large reduction in *R* for the older age groups during the vaccination roll out as case numbers are suppressed. Infection preferentially shifted to the unvaccinated age-groups, and our national R-number is weighted across subgroups by cases, not population. So the national *R* is dominated by the younger population.

Furthermore, *R* represents the rate of increase in infections, not the total numbers. Thus it is affected only by the rate of increase of vaccination, not the total numbers.

We see that *R* in the older age groups in July rebounded to the national average once almost everyone in those groups is vaccinated. This similar *R* across age groups implies that they are mixing. Given the relative prevalences, it represents infection of the older ages by the younger, unvaccinated population, rather than transmission within one age-band. The case numbers in the older age groups remain low thanks to the strong suppression of *R* during the vaccine roll-out.

### 4.3 Detection of Events

It is possible to detect individual events in the data, and test correlations by investigating appropriate subdivision. For example, the peak in December associated with the alpha variant can be seen to occur earliest in the South East and later further north, consistent with its believed origins in Kent. Conversely, the peak associated with the delta variant appears first in the North West, then almost simultaneously everywhere else, suggesting multiple importations rather than geographical spread.

Increases correlated with reopening of schools can be seen to occur first in the youngest age groups and the typical age of parents, again strongly suggestion of causation. Furthermore, the peak in July which has been associated with sporting events such as EURO2021 can be seen to be initially driven by men, spreading subsequently to women.

### 4.4 Features beyond R

The WSS approach can be applied not only to I(t) and C(t), but also to any other quantity, for example hospitalization or deathrate. Unlike conventional ODE-driven compartment models, WSS incorporates a delay moving from one compartment to the next: thus cases are related to deaths via a generalization of Eq,

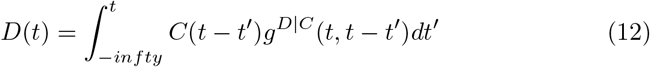

Where *g*^*D*|*C*^ is the probability of death at time *t* given a case reported at time *t*′. Note that the forwards projection avoid the entropy-decrease problem discussed in section 4, correctly predicting that sharp peaks in *C*(*t*) will lead to broader peaks in *D*(*t*).

We write *g*^*D*|*C*^(*t, t* − *t*′) as a function of two variables. The *t* − *t*′ dependence represents the trajectory of the illness from infection to death: this has been determined in clinical studies. The *t* dependence represents changes in disease severity over time. Disentanglng these, we can write:

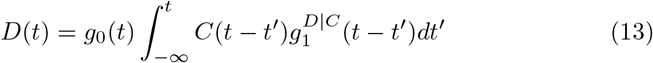

This *g*_0_ is a time-dependent case-fatality ratio, and can be plotted as a ratio of observed to predicted deaths. 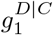 provides the shift forward in time from case to death: it is fitted using a normalised Gamma distribution. *g*_0_ is extremely dependent on agegroup: we use separate functions for each five-year age band. (*D*_*obs*_(*t*)*/D*_*W SS*_(*t*)). Fig.8 provides a powerful image of the lethality of the epidemic. There are three salient features.

The alpha variant is accompanied by a pronounced increase in CFR through December, plateauing once alpha is ubiquitous. The sharp decline in 2021, and the onset of the effect in oldest age group first, can be associated with the effect of the vaccine in causing milder infections. The age-dependence of CFR is so pronounced that for under-45s (not shown) statistics are too poor for reasonable analysis.

A discernable blip in the drop of CFR during May 2021 could be associated with the arrival of the delta variant (Fig 8).

**Figure 8:**
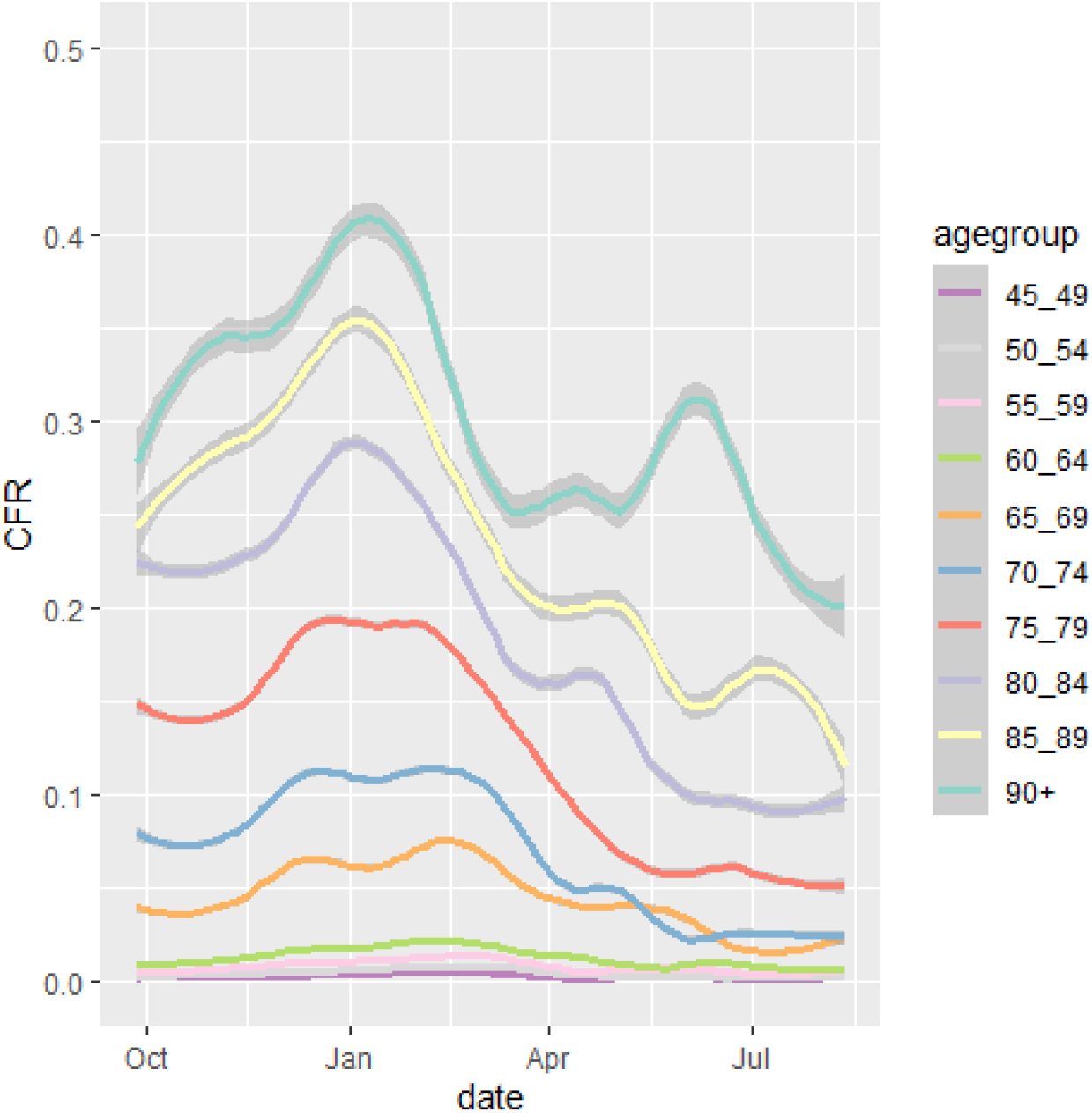
Case Fatality ratios, plotted as Deaths per Case from WSS model, by age group. Lines are a weighted smoothed fit to the data. CFR graphs for under 45s are excluded as they are so low. Shading shows uncertainty introduced by smoothing day to day variations, excluding errors on the mean from small number statistics in September 2020 and May 2021. The eye-catching peak for over 90s in June 2021 is probably a small-number effect, and it can be eliminated completely by combining 85-89 with 90+ age groups.

**Figure 9:**
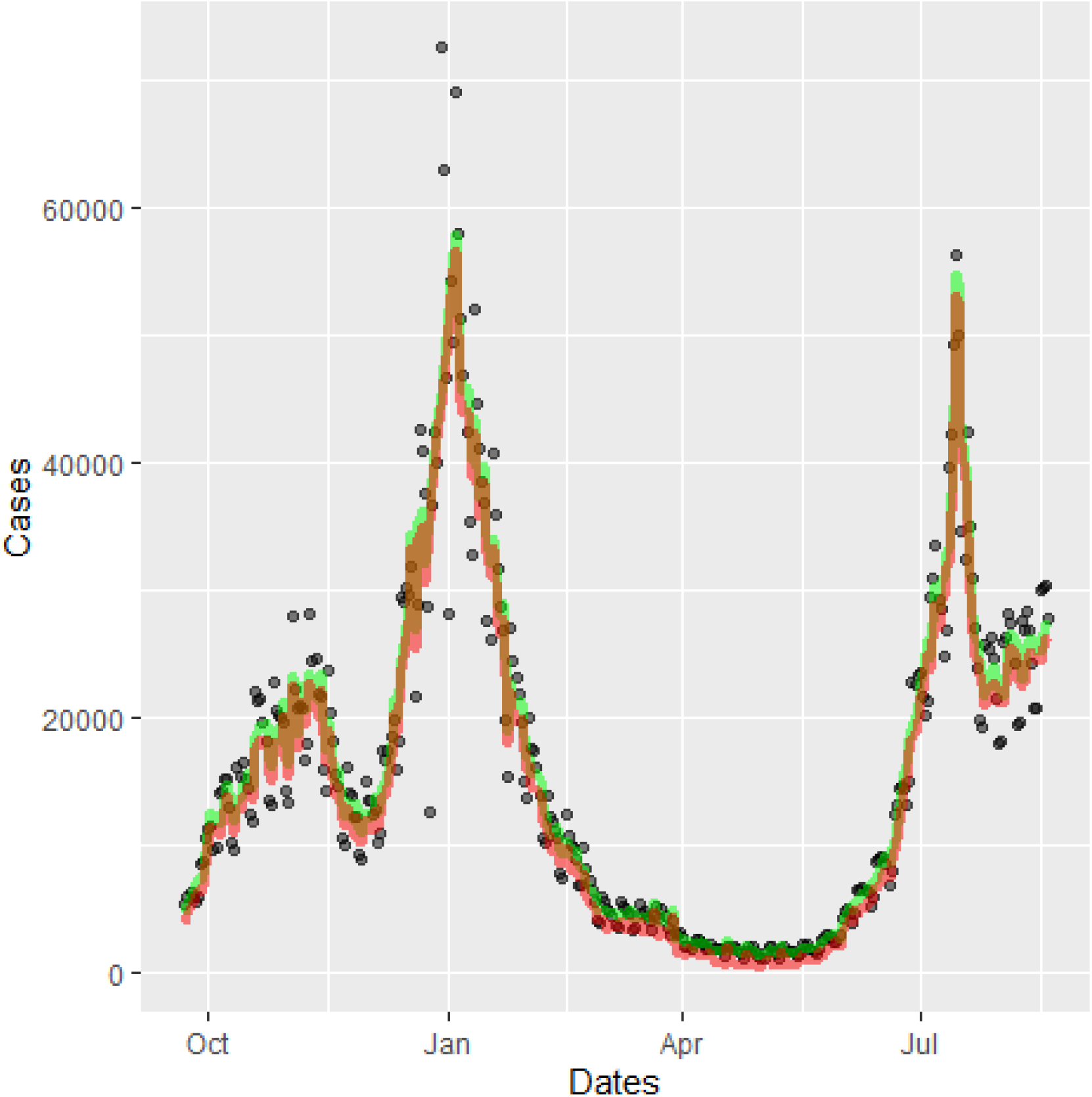
Effect of removing weekend and Christmas systematic errors: Positive first PCR test data as published (circles) [1],Weekend and Xmas Smoothed case data (green) and Corrected for 0.4% false positives (red)[21].

As well as age-related factors, WSS enables us to discern differences in CFR across different geographical regions. This indicates a strong north-south divide, as has been discussed in previous work[6, 21]. CFR is significantly lower in the south except for a short window when alpha variant was more prevalent there.

## 5 Conclusions

The published *R*-number from SPI-M can be predicted some 16 days in advance of publication by statistical analysis of the publicly available case data using our WSS code.

Our case-data estimates are themselves necessarily delayed by the time between infection and positive test, thus it is likely that the published values are around three weeks delayed from the actual spreading events. It is likely that the SPI-M modelling will provide more reliable estimate, however our WSS model appears to be adequate for making coarse policy-making decisions. For some applications, the earlier availability is likely to outweigh the loss of accuracy.

Case-number-based *R* smooths any sharp discontinuities in infection-based *R*. WSS is probably less reliable as a tool for analysing the effect of non-pharmaceutical interventions (NPIs) than models which incorporate infections explicitly and are parameterised using a Bayesian approach. However, the WSS *R* allows us to create a narrative of the second wave:

- Increase of cases through Sep-Oct, with *R* > 1.
- Sharp reduction of *R* with NPIs in November, rebounding as B1.117 (alpha) becomes established.
- Sharp drop of *R* at January lockdown (case data show a continuous drop, but this is consistent with a sharp drop in infections after Jan 6th, smoothed by variable incubation times).
- Steady rise in *R* throughout February-May, accelerating as B1.617 (delta) becomes established and restrictions were released.
- Sharp peak and drop in *R* in July, despite relaxation of restrictions.
- Rise in *R* in Scotland during August, not mirrored in England.

The WSS code also produces up-to-date Case-Fatality Ratios[21]. Analysis of these in Figure.8 shows a sharp decrease in the CFRs correlated with the vaccine roll out, showing that vaccination has a double-benefit of reducing infections and ameliorating the effects of Covid. The reduction in CFR is about 50%, and this has continued during the rise of the B1.617 variant. Correlation does not imply causation, but a protective effective of vaccination seems more likely than other possibilities consistent with the data, for example B1.617 being less deadly than B1.117.

We note that it may seem counterintuitive that *R* is increasing during the vaccination programme. This is because *R* derived from case data is not the average over the population but rather the average over those who are infected. Eliminating infection from a vaccinated subpopulation would mean reported *R* refers only to the unvaccinated population. Perhaps the most surprising outcome of this study is the excellent agreement of this simple method with far more detailed epidemiological models. This indicates that the case data currently being produced is sufficient to track the trajectory of the epidemic.

The R-number is well defined but unmeasureable in terms of who-infected-whom. It can be inferred from case date, however its relation to growth rate rests on the assumption of short term exponential growth with slowly varying R. This follows from a well-mixed ODE implementation of SIR or related models, whereas a lattice-based implementation SIR gives linear growth. These are limiting cases of a range of network models. The data from the UK coronavirus epidemic has features closer to the lattice-model end of the spectrum. The R-number has remained close to 1, with external shocks such as variants producing transient peaks in R of a few weeks’ duration before returning to 1. This happened both with a lockdown in January, and without one in July. Similarly, the epidemics is more reliably reproduced from R-numbers derived from smoothed cases, rather than smoothing the R-number itself. This indicates that short term fluctuations in case data are additive rather than multiplicative implying medium term linear growth, rather than exponential growth.

The effects of lockdowns etc. in reducing cases and suppressing spread are significant in all cases - in a well mixed model this manifests in a lowered herd immunity threshhold, it the lattice models as a slower moving wavefront. Long distance travel bans have the effect of reducing long range connections, making the network more lattice-like.

The lattice model indicates that an initial value of *R*_0_ above 2 is required to generate a sustained epidemic, as opposed to 1 for a well-mixed model. However, if the disease spreads as a wave it generates slightly higher total case numbers than the well-mixed case. We note that an *observed R*(*t*) = 1 value in consistent with a much higher *R*_0_, and that significant reduction of *R*_0_ may have little effect on *R*(*t*): individuals at the wavefront can only become infected once, even when a high *R*_0_ implies they may have several encounters which could lead to infection.

Medium term epidemic predictions for hospital occupation, ICU demand and deaths are extremely sensitive to assumptions regarding *R*(*t*), which most interventions target *R*_0_. It is therefore crucial to understand the relationship between them. As deduced the UK case data, R has remained close to 1 with occasional excursions producing short-lived transients. The alpha variant, originally detected in Kent, spread geographically from south to north in the period of a couple of months. It appears that the coronavirus is spreading is a network dominated by localised interactions.

## Data Availability

Code for this work is available in github
All data is collected from publicly available websites

https://github.com/gjackland/WSS

## 6 Acknowledgements

GJA and MA were funded by UKRI under grant ST/V00221X/1. We acknowledge support from the Royal Society RAMP initiative and thank Rowland Kao for helpful comments.

## 7 Appendix

## Footnotes

1 In which case it is equivalent to growth rate.

2 e.g. It could be modelled as a reaction-diffusion process in two dimensions, for which cases grow quadratically in time

3 Another term used ambiguously in the literature

4 Changes in calculated *R* will exhibit some delay from the exponential decay of *R <* 1, and local suppression measures will speed this decay.

5 This assumes independent infection events, if superspreading events are significant the noise will be larger, although some smoothing is already provided by the distribution of times from infection to test

6 The reported number of cases for England is approximately 1% higher than the sum of the regions, this is because of cases which cannot be assigned to any region

7 This is the obvious stochastic cellular automaton generalization of SIR. We first coded it for the DAP computer installed at EPCC 1984, when it was already a well-established demonstration application for SIMD archetecture

8 which fail if the target is already in state I or R

9 https://www.gov.uk/government/groups/scientific-pandemic-influenza-subgroup-on-modelling

10 https://www.gov.uk/government/publications/reproduction-number-r-and-growth-rate-methodology/reproduction-number-r-and-growth-rate-methodology

